# Decline in mortality among hospitalised covid-19 patients in Sweden: a nationwide observational study

**DOI:** 10.1101/2020.10.27.20220061

**Authors:** Kristoffer Strålin, Erik Wahlström, Sten Walther, Anna M Bennet-Bark, Mona Heurgren, Thomas Lindén, Johanna Holm, Håkan Hanberger

## Abstract

**OBJECTIVE:** It is important to know if mortality among hospitalised covid-19 patients has changed as the pandemic has progressed. The aim of this study was to describe the dynamics of mortality among patients hospitalised for covid-19 in a nationwide study.

**DESIGN:** Nationwide observational cohort study of all patients hospitalised in Sweden 1 March to 30 June 2020 with SARS-CoV-2 RNA positivity 14 days before to 5 days after admission, and a discharge code for covid-19.

**SETTING:** All hospitals in Sweden.

**PARTICIPANTS:** 15 761 hospitalised patients with covid-19, with data compiled by the Swedish National Board of Health and Welfare.

**MAIN OUTCOME MEASURES:** Outcome was 60-day all-cause mortality. Patients were stratified according to month of hospital admission. Poisson regression was used to estimate the relative risk of death by month of admission, adjusting for pre-existing conditions, age, sex, care dependency, and severity of illness (Simplified Acute Physiology, version 3), for patients in intensive care units (ICU).

**RESULTS:** The overall 60-day mortality was 17.8% (95% confidence interval (CI), 17.2% to 18.4%), and it decreased from 24.7% (95% CI, 23.0% to 26.5%) in March to 13.3% (95% CI, 12.1% to 14.7%) in June. Adjusted relative risk (RR) of death was 0.56 (95% CI, 0.51 to 0.63) for June, using March as reference. Corresponding RR for patients not admitted to ICU and those admitted to ICU were 0.60 (95% CI, 0.53 to 0.67) and 0.61 (95% CI, 0.48 to 0.79), respectively. The proportion of patients admitted to ICU decreased from 19.5% (95% CI, 17.9% to 21.0%) in the March cohort to 11.0% (95% CI, 9.9% to 12.2%) in the June cohort.

**CONCLUSIONS:** There was a gradual decline in mortality from March to June 2020 in Swedish hospitalised covid-19 patients, which was independent of pre-existing conditions, age, and sex. Future research is needed to explain the reasons for this decline. The changing covid-19 mortality should be taken into account when management and results of studies from the first pandemic wave are evaluated.

## Introduction

The Coronavirus Disease 2019 (covid-19) pandemic has put enormous pressure on the healthcare system in general and on hospitals in particular, despite extensive interventions to reduce spread of the coronavirus SARS-CoV-2.^1^ Patients admitted for covid-19 have been reported to have mortality fractions of ≥20% overall^2-5^ and of >34% among patients admitted to an intensive care unit (ICU).^2,6-8^ The proportion of patients requiring ICU admission is reported to be 17-32%.^2,9-11^

Most studies on covid-19 mortality have included patients admitted between February and April 2020, i.e. early in the covid-19 pandemic.^1^ Since the outbreak of the pandemic, there has been a gradual and substantial increase in our understanding of covid-19. This may have contributed to the improved survival that has been noted among ICU-treated covid-19 patients. In an ICU study of covid-19 patients in England, Wales and Northern Ireland by Doidge et al.,^8^ a decline in 28-day mortality was noted from 43.5% before the peak of the first pandemic wave to 34.3% after the peak. A meta-analysis of Armstrong et al.^12^ showed lower ICU mortalities in studies published April-May than in those published January-March. However, a large proportion of hospitalised covid-19 patients are never admitted to ICU, and to our knowledge it has not been clarified if mortality has changed among non-ICU treated hospitalised patients. Karagiannidis et al.^5^ reported no considerable change in mortality over time in a large German cohort of unselected hospitalised covid-19 patients.

The present Swedish study was undertaken to examine whether mortality has changed with time in a nationwide cohort of hospitalised covid-19 patients. The specific aim was to evaluate nationwide 60-day mortality separately for non-ICU treated and ICU treated patients, during the first 4 months of the pandemic, using data compiled by the Swedish National Board of Health and Welfare (NBHW).

## Methods

### Study design and setting

Nationwide observational cohort study on SARS-CoV-2-positive individuals treated for covid-19 in Swedish hospitals.

### Participants

The study population was derived from cross-linked national population-based registers using the unique personal identity number assigned to each Swedish resident at birth or on immigration to Sweden.^13^

From the National Patient Register (NPR), held by the NBHW, all patients admitted to hospitals in Sweden between 1 March and 30 June, 2020 were identified, see flowchart in fig 1.Cross-linking with the Swedish reporting system for notifiable infectious diseases (SmiNet)^14^ provided data on SARS-CoV-2 PCR test results. Hospitalised patients with a PCR test result positive for SARS-CoV-2 RNA 14 days before to 5 days after admission were then identified. Finally, those with a discharge code of covid-19, i.e. U07.1 according to the *10*^*th*^ *International Statistical Classification of Diseases* (ICD-10), (n=15 761) constituted the study population of the present study.

**Fig 1.**
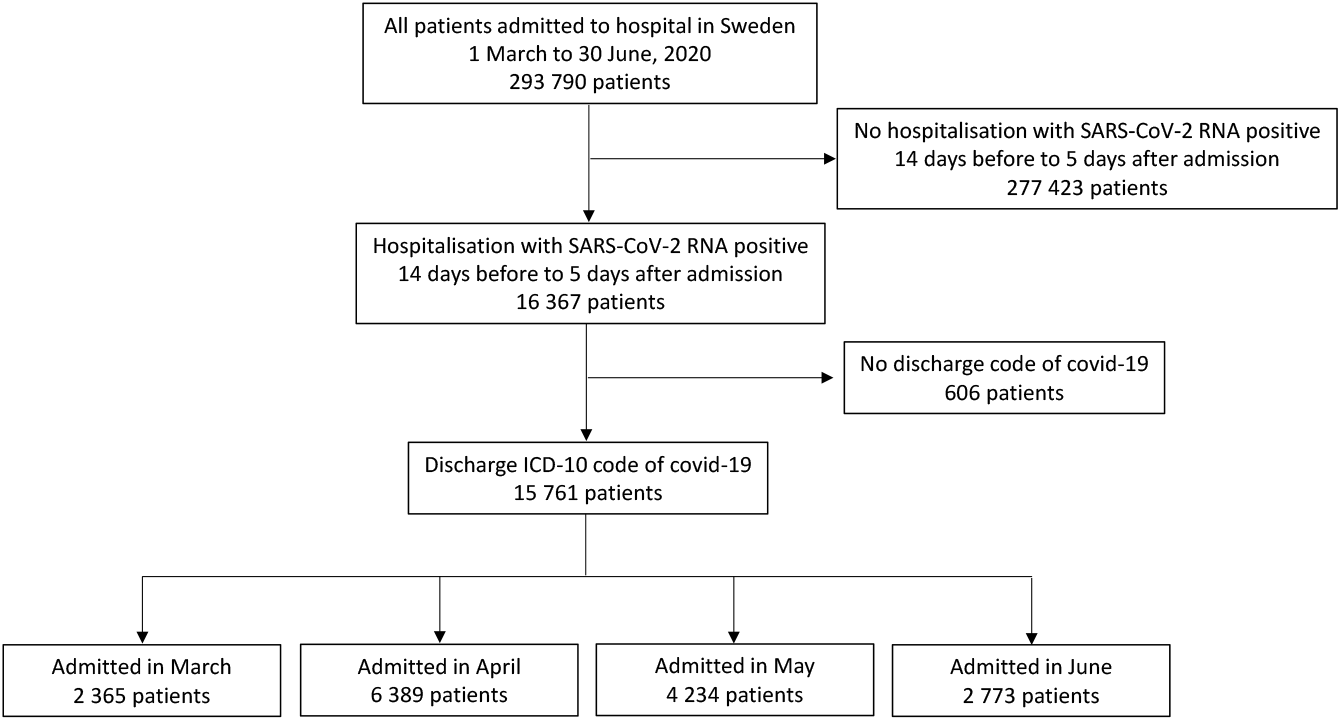
Flow chart of study inclusion: Patients hospitalised for covid-19 in Sweden 1 March – 30 June, 2020

### Outcome and covariate data

Primary outcome was 60-day mortality, defined as death from any cause within 60 days of index hospital admission. Sixty days was considered to be a reasonable follow-up time since very few patients remain in hospital for longer periods, but patients in ICU often remain hospitalised beyond 30 days.^5^ Date of death was obtained from the Swedish Cause of Death Register^15^ (held by NBHW). Since the beginning of the covid-19 pandemic, the register has been updated daily with all dates of death, as reported to the Swedish Tax Agency (mandatory reporting by law), resulting in no loss to follow-up.

ICU episodes were identified from the Swedish Intensive Care Registry, a national quality register for intensive care in Sweden including 83 of 84 ICUs in the country.^16^ Since the outbreak of the pandemic the register has had complete coverage of all ICU treated covid-19 patients in Sweden.

In the present study, patients were stratified according to month of admission, in line with the Swedish epidemiological curve of new admission for covid-19 (fig 2), i.e. rapid increase in admission rate in March, relatively stable high admission rate in April, relatively stable moderately high admission rate in May, and decreasing admission rate in June. The date of index admission for each patient was defined as the admission date of the first hospital care episode that fitted the inclusion criteria.

**Fig 2.**
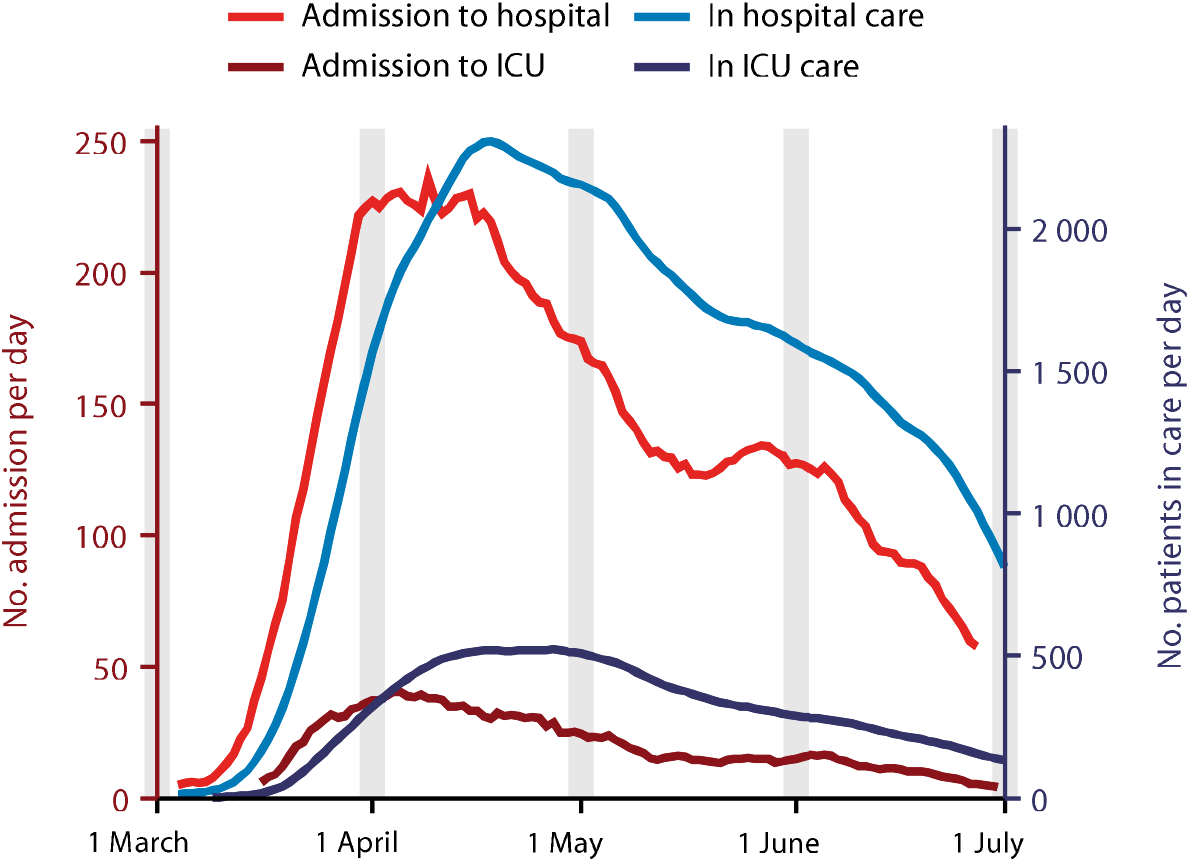
Timeline of patients admitted to, and in care, in hospitals for covid-19 in Sweden during the study period. Shown are number of patients admitted per day into hospital (by index admission date), and into ICU specifically (by ICU admission date) on left Y-axis; number of patients in care per day in hospital, and in ICU specifically on right Y-axis

To account for patient transfer within or between hospitals, resulting in multiple entries in the NPR, duration of hospital stay was defined as number of days between index admission date and last discharge date in cases with sequential admissions. A sequential admission was defined as a readmission occurring within 1 day of the previous one, starting with the index admission. Duration of hospital stay in days was compared using the outcome discharge status (alive/deceased) rather than 60-day mortality, to avoid truncating one group at 60 days by default.

Comorbidities were defined using discharge diagnoses for the last five years and/or prescribed drugs dispensed during the year preceding index admission. A 30-day wash-out window prior to index admission date was applied. Codes defining each comorbidity are provided in table E1. In addition, Charlson Comorbidity Index (CCI) was calculated as described in table E2.

Information on discharge diagnoses were identified through the NPR, which contains information on all reported cases of inpatient care and/or visits to a physician at a specialised outpatient clinic in Sweden. The validity of the register is generally high with positive predictive values of 85% to 95% for most diagnoses in validation studies.^17^

Data on drugs dispensed were obtained from the Swedish Prescribed Drug Register (held by NBHW), which contains all prescribed and pharmacy-dispensed medicines in the community classified according to the Anatomic Therapeutic Chemical Classification System (ATC). It has almost complete coverage (data missing <1%).^18^

Information on care dependency (nursing home resident or homecare) prior to admission was obtained from the Care and Social Services for the Elderly and for Persons with Impairments Register (held by NBHW). Date and country of birth were obtained from the NPR.

Data on Simplified Acute Physiology, version 3 (SAPS3), oxygenation index (PaO2/FiO2), treatments and procedures on the ICU were obtained from the Swedish Intensive Care Registry. For SAPS3 and PaO2/FiO2, information from the first admission only was used. If a patient had several ICU admissions, treatment information was retrieved for all episodes of ICU care for covid-19.

### Statistical analyses

Baseline patient characteristics were tabulated by month of admission, ICU treatment and survival outcome. Confidence intervals (CI) were calculated for proportions using the Wilson Score interval and expressed as percentages.

Crude survival curves with 95% Hall-Wellner confidence bands were estimated using the Kaplan-Meier estimator, plotted by month of admission with all-cause mortality as event. Follow-up was truncated at 60 days after index admission. The log-rank test was used to test for differences in survival between groups, with a two-sided alpha of 0.05.

Furthermore, percentage mortalities were plotted by month of admission, overall, by age groups (<60, 60-69, 70-79, ≥80), sex, and healthcare region.

Incident number of covid-19 patients admitted to hospital per day was plotted by index admission date for the whole patient cohort as well as separately for ICU, using the first occurring ICU admission date for a patient. The number of covid-19 patients residing in hospital care per day was also calculated, again showing one curve for the overall cohort and one for the ICU treated cohort separately. Lines were smoothed using a seven-day rolling average, based on three days prior-three days after admission.

Relative risk (RR) with 95% CI was estimated in multivariable analysis for the outcome 60-day mortality with month of hospitalisation as exposure of interest, using modified Poisson regression models.^19^ Estimates of RR were obtained from three models: Model 1 adjusted for age (continuous, both a linear and quadratic term), sex (male/female), CCI (categorical, 0, 1-2, 3-4, 5+), care dependency (nursing home, homecare, neither), country of birth (Sweden/other), and healthcare region (North, Uppsala-Örebro, Stockholm-Gotland, Southeast, West, and South). Model 2 additionally included an interaction term between month of admission and healthcare region. Model 3 estimated age as a categorical variable (<60, 60-69, 70-79, ≥80) for increased interpretability, and included an interaction term between month of admission and age group.

Model 1 was also stratified between ICU admission or not during the hospital episode. In cases of ICU care, the SAPS3 score (continuous) for the first ICU episode was added, creating Model 4.

In all regression models, missing data were handled by complete case analysis. Data were complete on all variables except for country of birth and CCI (5% missing) and SAPS3 (<1% missing). Sensitivity analyses were performed comparing adjusted RR when imputing missing country of birth to ‘Sweden’ or ‘Outside Sweden’, and when imputing missing CCI to extreme values ‘0’ or ‘5+’.

All data management and statistical analyses were performed using SAS Software SAS Enterprise guide v7.15, SAS Institute Inc., Cary, NC.

### Patient involvement

Patients or the public were not involved in the design, conduct, reporting, or dissemination plans of our research.

### Ethics and Reporting

Ethical approval for the study was obtained from the Swedish Ethics Review Authority, Uppsala (Dnr 2020-04278). The study conforms to the Reporting of Observational Studies in Epidemiology (STROBE) statement.^20^

## Results

### Study Population

Altogether, 15 761 patients admitted for covid-19 in Swedish hospitals were studied, median age 64 years (interquartile range, 51 to 78 years), 57.5% men and 42.5% women. Fig 2 shows the number of new hospital admissions and the total numbers of in-patients on each time point. The peak numbers of patients admitted and in hospital care occurred during April. The proportion of patients admitted to ICU was 14.4% (95% CI, 13.9% to 15.0%).

Patient characteristics, stratified according to month of admission, are shown in table 1. The proportion of men decreased from 60.1% (95% CI, 58.1% to 62.1%) in March to 54.2% (95% CI, 52.4% to 56.1%) in June. Age distribution changed over time with the number of patients < 40 years and > 90 years increasing between March and June. The proportion of patients with a CCI of zero was 58.1% (95% CI, 56.1% to 60.1%) in March and 63.7% (95% CI, 61.9% to 65.5%) in June. In March 42.2% (95% CI, 40.2% to 44.2%) of patients were born outside Sweden compared to 33.7% (95% CI 32.0% to 35.5%) in June.

**Table 1.**
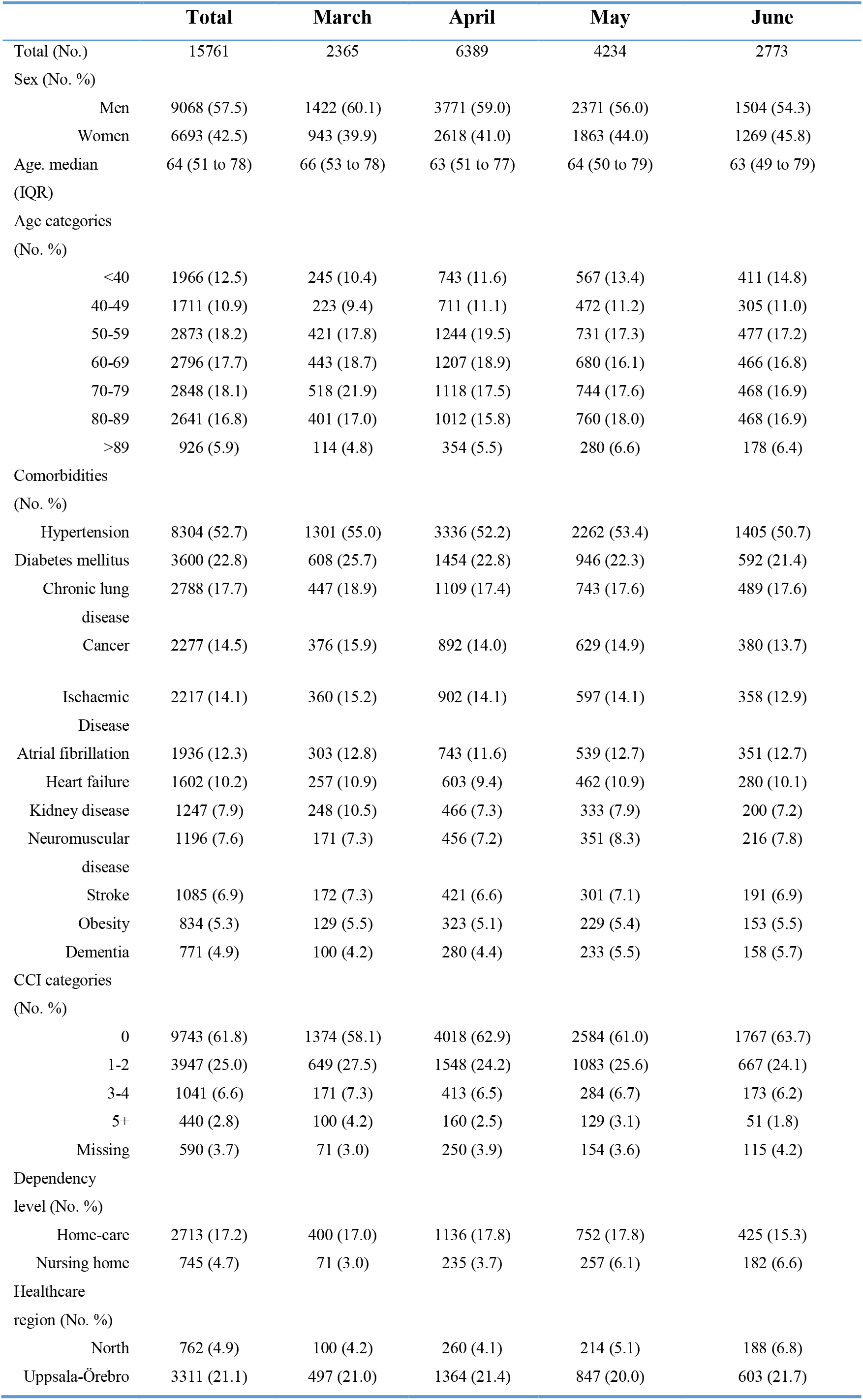
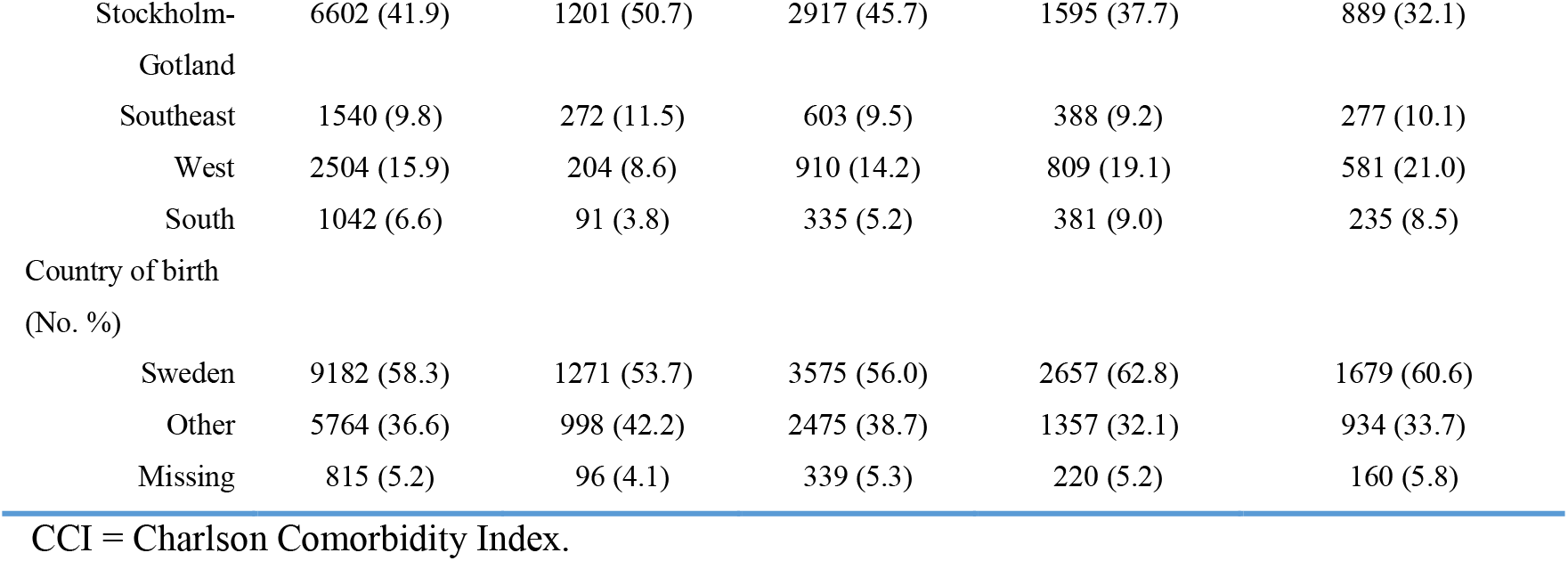
Baseline patient characteristics, total and stratified by month of admission

The duration of hospital stay is presented in table E3. The duration was clearly longer for patients admitted to ICU than for those not, but there was no clear change in duration over time.

### Overall mortality

The overall 60-day mortality was 17.8% (95% CI, 17.2% to 18.4%), it was 25.5% (95% CI, 23.7% to 27.3%) among patients admitted to ICU and 16.5% (95% CI, 15.8% to 17.1%) among patients not admitted to ICU. Table 2 shows patient characteristics for survivors and non-survivors at 60-day follow-up, stratified according to patients admitted or not admitted to ICU during hospital stay.

**Table 2.**
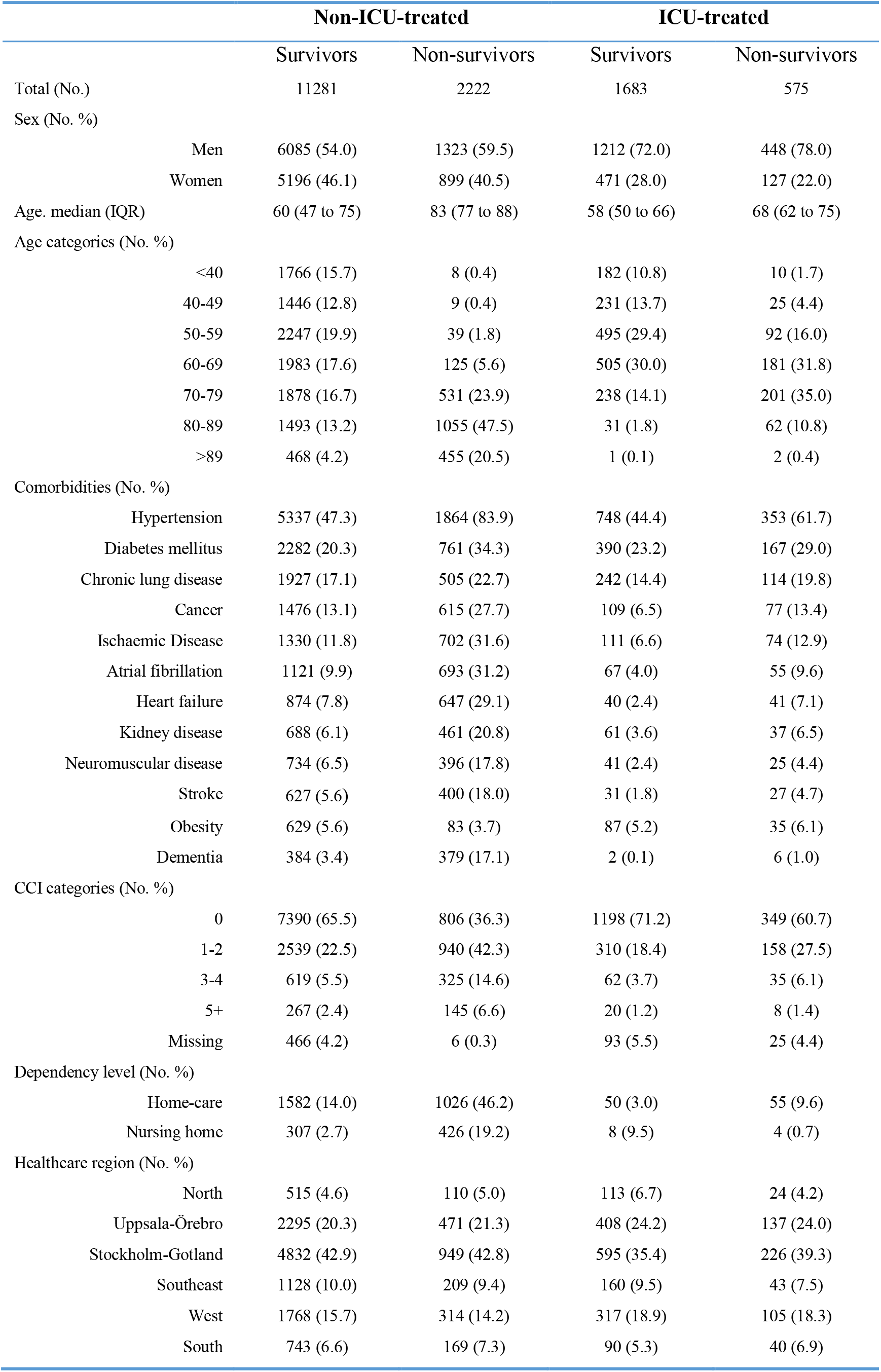
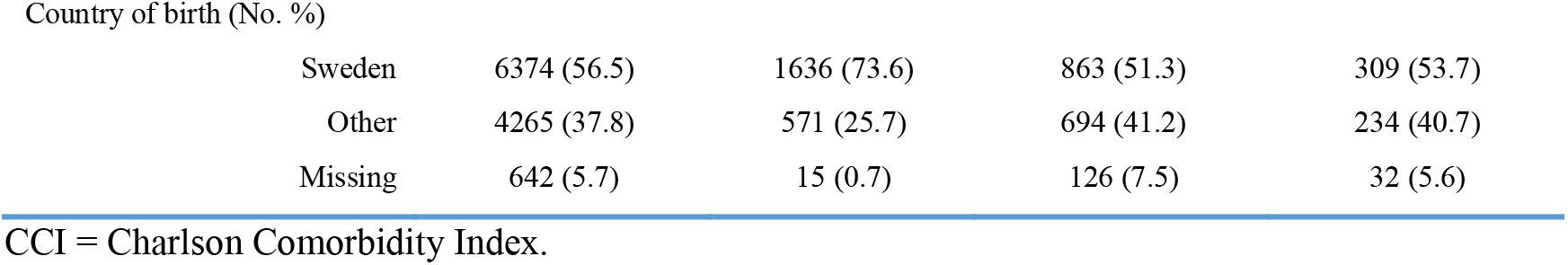
Baseline patient characteristics stratified by intensive care unit (ICU) treatment and survival outcome at 60-days follow-up

|  |  | Non-ICU-treated |  | ICU-treated |  |
| --- | --- | --- | --- | --- | --- |
|  |  | Survivors | Non-survivors | Survivors | Non-survivors |
| Total (No.) |  | 11281 | 2222 | 1683 | 575 |
| Sex (No. %) |  |  |  |  |  |
|  | Men | 6085 (54.0) | 1323 (59.5) | 1212 (72.0) | 448 (78.0) |
|  | Women | 5196 (46.1) | 899 (40.5) | 471 (28.0) | 127 (22.0) |
| Age, median (IQR) |  | 60 (47 to 75) | 83 (77 to 88) | 58 (50 to 66) | 68 (62 to 75) |
| Age categories (No. %) |  |  |  |  |  |
|  | <40 | 1766 (15.7) | 8 (0.4) | 182 (10.8) | 10 (1.7) |
|  | 40-49 | 1446 (12.8) | 9 (0.4) | 231 (13.7) | 25 (4.4) |
|  | 50-59 | 2247 (19.9) | 39 (1.8) | 495 (29.4) | 92 (16.0) |
|  | 60-69 | 1983 (17.6) | 125 (5.6) | 505 (30.0) | 181 (31.8) |
|  | 70-79 | 1878 (16.7) | 531 (23.9) | 238 (14.1) | 201 (35.0) |
|  | 80-89 | 1493 (13.2) | 1055 (47.5) | 31 (1.8) | 62 (10.8) |
|  | >89 | 468 (4.2) | 455 (20.5) | 1 (0.1) | 2 (0.4) |
| Comorbidities (No. %) |  |  |  |  |  |
|  | Hypertension | 5337 (47.3) | 1864 (83.9) | 748 (44.4) | 353 (61.7) |
|  | Diabetes mellitus | 2282 (20.3) | 761 (34.3) | 390 (23.2) | 167 (29.0) |
|  | Chronic lung disease | 1927 (17.1) | 505 (22.7) | 242 (14.4) | 114 (19.8) |
|  | Cancer | 1476 (13.1) | 615 (27.7) | 109 (6.5) | 77 (13.4) |
|  | Ischaemic Disease | 1330 (11.8) | 702 (31.6) | 111 (6.6) | 74 (12.9) |
|  | Atrial fibrillation | 1121 (9.9) | 693 (31.2) | 67 (4.0) | 55 (9.6) |
|  | Heart failure | 874 (7.8) | 647 (29.1) | 40 (2.4) | 41 (7.1) |
|  | Kidney disease | 688 (6.1) | 461 (20.8) | 61 (3.6) | 37 (6.5) |
|  | Neuromuscular disease | 734 (6.5) | 396 (17.8) | 41 (2.4) | 25 (4.4) |
|  | Stroke | 627 (5.6) | 400 (18.0) | 31 (1.8) | 27 (4.7) |
|  | Obesity | 629 (5.6) | 83 (3.7) | 87 (5.2) | 35 (6.1) |
|  | Dementia | 384 (3.4) | 379 (17.1) | 2 (0.1) | 6 (1.0) |
| CCI categories (No. %) |  |  |  |  |  |
|  | 0 | 7390 (65.5) | 806 (36.3) | 1198 (71.2) | 349 (60.7) |
|  | 1-2 | 2539 (22.5) | 940 (42.3) | 310 (18.4) | 158 (27.5) |
|  | 3-4 | 619 (5.5) | 325 (14.6) | 62 (3.7) | 35 (6.1) |
|  | 5+ | 267 (2.4) | 145 (6.6) | 20 (1.2) | 8 (1.4) |
|  | Missing | 466 (4.2) | 6 (0.3) | 93 (5.5) | 25 (4.4) |
| Dependency level (No. %) |  |  |  |  |  |
|  | Home-care | 1582 (14.0) | 1026 (46.2) | 50 (3.0) | 55 (9.6) |
|  | Nursing home | 307 (2.7) | 426 (19.2) | 8 (0.5) | 4 (0.7) |
| Healthcare region (No. %) |  |  |  |  |  |
|  | North | 515 (4.6) | 110 (5.0) | 113 (6.7) | 24 (4.2) |
|  | Uppsala-Örebro | 2295 (20.3) | 471 (21.3) | 408 (24.2) | 137 (24.0) |
|  | Stockholm-Gotland | 4832 (42.9) | 949 (42.8) | 595 (35.4) | 226 (39.3) |
|  | Southeast | 1128 (10.0) | 209 (9.4) | 160 (9.5) | 43 (7.5) |
|  | West | 1768 (15.7) | 314 (14.2) | 317 (18.9) | 105 (18.3) |
|  | South | 743 (6.6) | 169 (7.3) | 90 (5.3) | 40 (6.9) |

Country of birth (No. %)
|  |  |  |  |  |
| --- | --- | --- | --- | --- |
| Sweden | 6374 (56.5) | 1636 (73.6) | 863 (51.3) | 309 (53.7) |
| Other | 4265 (37.8) | 571 (25.7) | 694 (41.2) | 234 (40.7) |
| Missing | 642 (5.7) | 15 (0.7) | 126 (7.5) | 32 (5.6) |

CCI = Charlson Comorbidity Index.

### Changes in Mortality Over Time

Fig 3 shows that the overall 60-day mortality decreased from 24.7% (95% CI, 23.0% to 26.5%) in March to 13.3% (95% CI, 12.1% to 14.7%) in June. Likewise, it decreased from 36.1% (95% CI, 31.8% to 40.6%) to 20.1% (95% CI, 16.0% to 24.9%) for patients admitted to ICU, and from 21.9% (95% CI, 20.1% to 23.9%) to 12.5% (95% CI, 11.3% to 13.9%) for patients not admitted to ICU.

**Fig 3.**
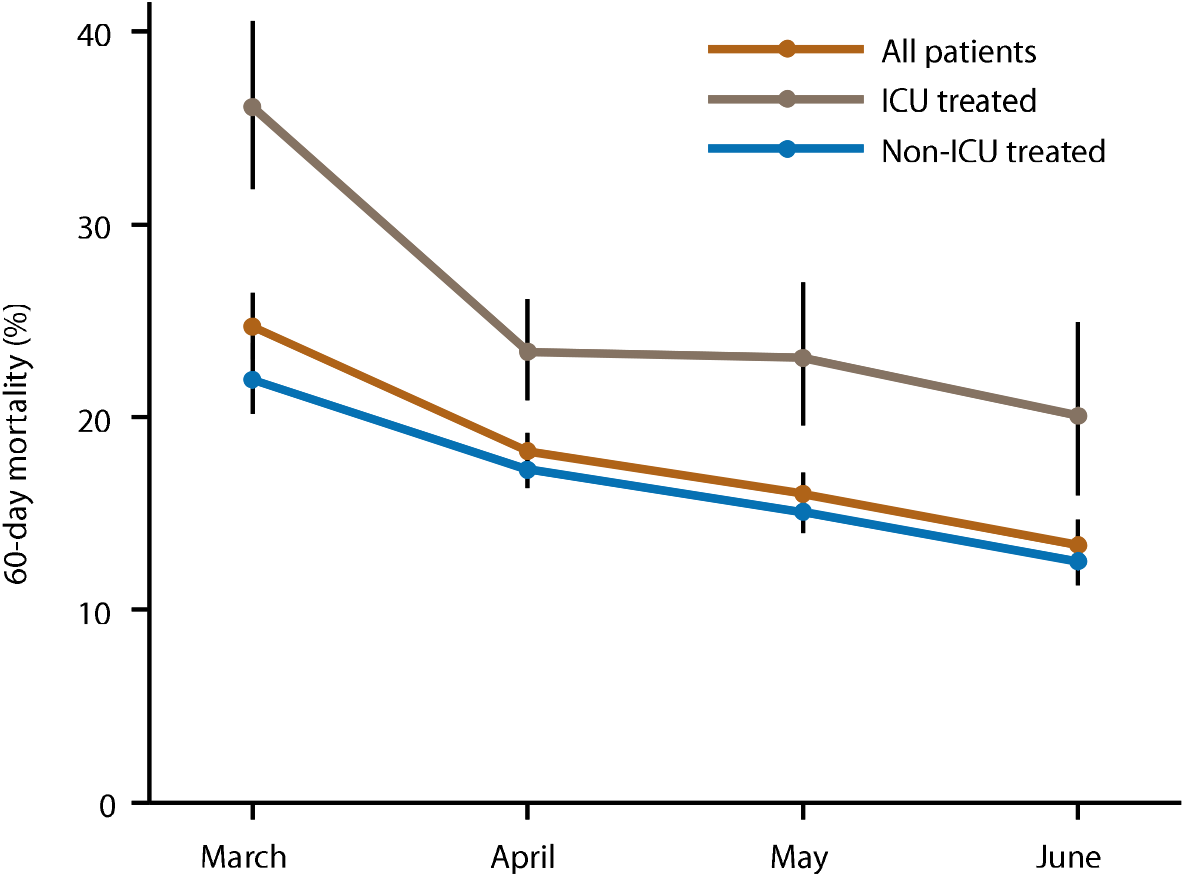
60-day mortality related to month of hospital admission. Proportions with 95% confidence intervals are shown per month of admission, total and separately for ICU- and non-ICU treated patients

Kaplan-Meier analysis confirmed mortality differences according to month of admission, with a log-rank p-value <0.001 for both non-ICU and ICU treated patients (fig 4). The survival curves showed a higher initial mortality rate for the March cohort than for the other months of admission, most clearly so for those admitted to ICU.

**Fig 4.**
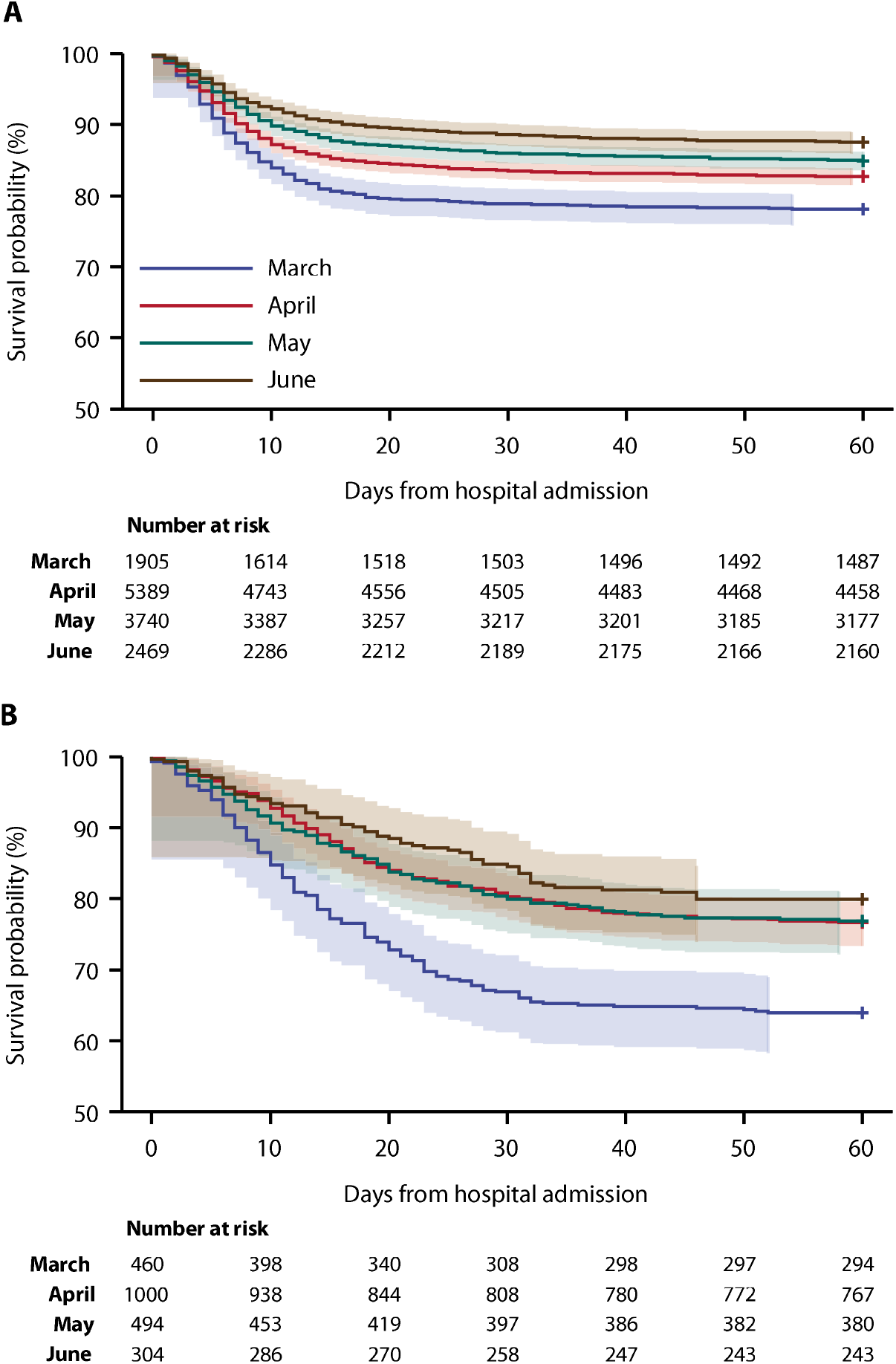
Kaplan-Meier estimator survival curves of patients not treated in ICU (A) and treated in ICU (B), according to month of hospital admission. Both (A) and (B) had log-rank p-value <0.001

In multivariable regression analyses, the decrease in mortality remained throughout the study period with an adjusted relative risk (RR) of 0.56 (95% CI, 0.51 to 0.63) in June, using March as reference. Adjusted mortality, with March as reference, decreased in patients not treated on ICU with RR 0.60 (95% CI, 0.53 to 0.67), and in patients treated on ICU with RR 0.61 (95% CI, 0.48 to 0.79) (fig 5, fig E1-E3). Sensitivity analysis showed no impact on results when imputing missing data on country of birth as “Sweden or outside Sweden” and with CCI as either of the two extreme values.

**Fig 5.**
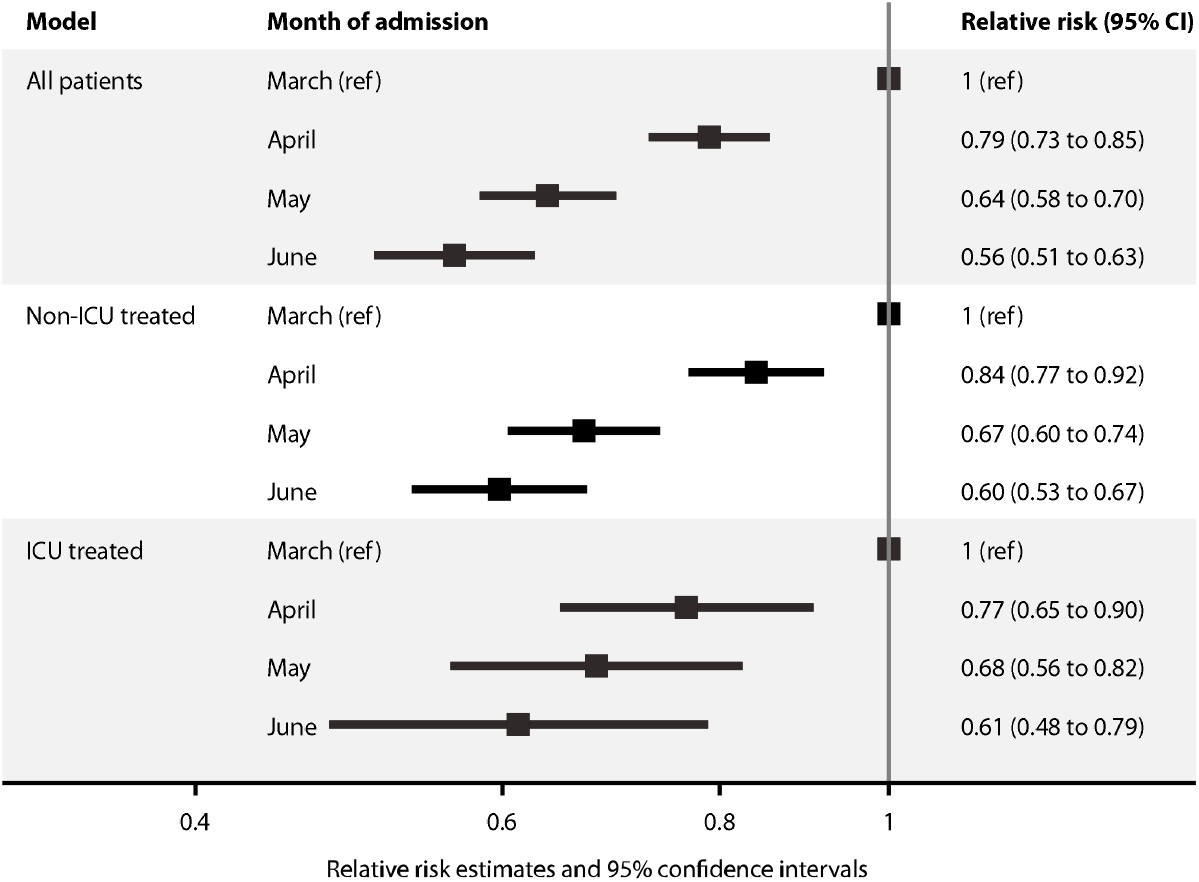
Relative risks with 95% confidence intervals for all-cause death within 60 days from hospital admission, for; (upper panel) All hospitalised patients, model adjusted for sex, age, Charlson Comorbidity Index, care dependency, country of birth and healthcare region; (middle panel) Patients not treated in ICU, model adjusted for sex, age, Charlson Comorbidity Index, care dependency, country of birth and healthcare region, (lower panel). ICU-treated patients, model adjusted for sex, age, Charlson Comorbidity Index, care dependency, country of birth, healthcare region and SAPS3 score at first ICU-admission

Fig E4-E6 show changes in mortality over time for age categories, sex, and healthcare regions. Tables E4-E5 show the relative risks from model 3 and 4, i.e. per month of admission separately by healthcare region and age respectively. There was a significant interaction both for age group and healthcare region (Joint test p<0.001). In the healthcare regions, the overall mortality in March ranged from 15.0% to 28.0%. The decline in mortality was most pronounced in the healthcare region with the highest initial mortality, i.e. Stockholm-Gotland, which also had the highest number of covid-19 cases/population (table E4 and E6, fig E6). In June, the overall mortality ranged from 10.3% to 15.3% in 5 healthcare regions, although it was 19.1% in one healthcare region (North).

### Change in variables over time among patients admitted to ICU

As shown in fig 6, the proportion of patients admitted to ICU decreased from 19.5% (95% CI, 17.9% to 21.0%) in the March cohort to 11.0% (95% CI, 9.9% to 12.2%) in the June cohort, and the overall proportion receiving invasive mechanical ventilation decreased from 16.8% (95% CI, 15.4% to 18.4%) to 6.4% (95% CI, 5.6% to 7.4%). Table 3 presents characteristics over time of patients admitted to ICU. The proportion of ICU patients receiving invasive mechanical ventilation decreased from 86.5% to 58.6% and the proportion receiving dialysis decreased from 22.8% to 13.8%, while the proportion of patients treated in the prone position increased in April and May compared to March. However, SAPS3 and PaO2/FiO2 on ICU admission remained unchanged during the study period.

**Table 3.** Characteristics of the patient population treated on intensive care units (ICU), by month of hospital admission and survival outcome at 60 days follow-up

|  | **March** | **April** | **May** | **June** | **Survivors** | **Non-survivors** |
| --- | --- | --- | --- | --- | --- | --- |
| **Total ICU population** | 460 | 1000 | 494 | 304 | 1683 | 575 |
| **Hospital days pre-ICU median (IQR)** | 1 (0 to 3) | 1 (0 to 3) | 1 (0 to 3) | 1 (0 to 3) | 1 (0 to 3) | 1 (0 to 3) |
| **Days on ICU, median (IQR)** | 13 (7 to 21) | 14 (7 to 24) | 13 (6 to 23) | 11 (4 to 21) | 13 (6 to 23) | 12 (6 to 22) |
| **SAPS3 mean (SD)** | 54.5 (10.2) | 52.9 (9.7) | 54.6 (10.6) | 54.3 (10.9) | 51.7 (8.9) | 59.7 (11.1) |
| **PaO2/FiO2 mean (SD) kPa** | 17.4 (13.7) | 16.1 (17.3) | 16.6 (13.7) | 15.7 (11.6) | 17.0 (16.7) | 14.9 (9.9) |
| **Invasive mechanical ventilation (No. % )** | 398 (86.5) | 761 (76.1) | 342 (69.2) | 178 (58.6) | 1202 (71.4) | 477 (83.0) |
| **Renal dialysis (No. % )** | 105 (22.8) | 181 (18.1) | 87 (17.6) | 42 (13.8) | 238 (14.1) | 177 (30.8) |
| **Prone position (No. % )** | 159 (34.6) | 489 (48.9) | 253 (51.2) | 135 (44.4) | 732 (43.5) | 304 (53.0) |
| **Corticosteroids (No. % )** | 24 (5.2) | 73 (7.3) | 75 (15.2) | 85 (27.9) | 185 (11.0) | 72 (12.5) |
| **Tocilizumab (No. % )** | 23 (5.0) | 17 (1.7) | 17 (3.4) | 4 (1.3) | 48 (2.9) | 13 (2.3) |
| **Remdesivir (No. % )** | 0 (0) | 9 (0.9) | 7 (1.4) | 5 (1.6) | 17 (1.0) | 4 (0.7) |

**Fig 6.**
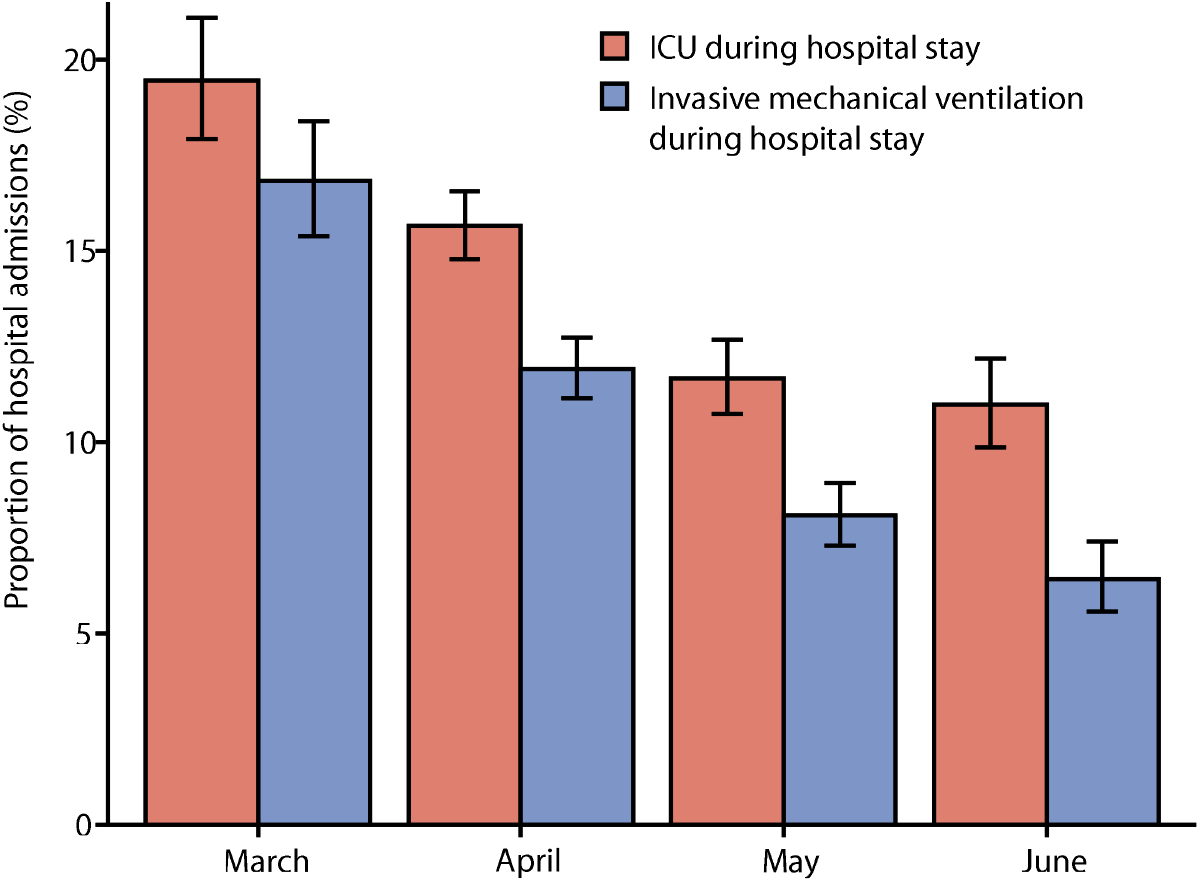
Proportion of patients treated in ICU and receiving invasive mechanical ventilation during the hospital stay, according to month of hospital admission. Shown are proportions with 95% confidence intervals

## Discussion

The present nationwide study on patients hospitalised for covid-19 in Sweden showed a distinct gradual decline in 60-day mortality between March and June both for non-ICU treated and ICU treated patients. The results were stable after adjustment for pre-existing conditions, age, sex, level of care dependency, country of birth, healthcare region, and SAPS3 (ICU treated patients).

The mortalities of 17.9% among hospitalised covid-19 patients overall and 25.5% among ICU treated covid-19 patients were low compared to reported mortalities from other countries, i.e. ≥ 20%^2-5^ and > 34%,^2,6-8^ respectively. Reasons for differences in outcome are difficult to analyse since studies use different outcome definitions, provide different patient characteristics, and often include patients that are still treated in hospital. Importantly, the time period of inclusion is probably crucial for the overall mortality and for differences between studies, since the mortality has clearly been changing over time. In the present study, the overall 60-day mortality declined from 24.7% in March to 13.3% in June, it declined from 36.1% to 20.1% among ICU treated patients, and from 21.9% to 12.5% among non-ICU treated patients. However, as noted in fig E6, the mortality dynamics over time varied between the Swedish healthcare regions. The reasons for these variations are not known, but a number of factors may have contributed to those and to the overall decline in mortality.

First, improvements of management and care have probably been of great importance for the declining mortality. For instance, the present study shows that even though acute severity of illness (SAPS3) among ICU-treated patients remained unchanged over time, the proportion of patients managed in the prone position increased, and the proportion receiving invasive mechanical ventilation decreased (table 3).

During the study period the drug therapy of patients with covid-19 changed continuously. In March 2020, many covid-19 patients in Sweden received off-label treatment with chloroquine phosphate/hydroxychloroquine,^21^ until this use was stopped by the European Medicines Agency on April 1. However, this use may not have affected outcome, since a randomised control trial (RCT) by Horby et al.^22^ found that patients treated with hydroxychloroquine had similar mortality as those receiving standard of care treatment. In March and early April, publications showed that anticoagulant therapy with low molecular weight heparin (LMWH) was associated with a better prognosis^23^ and that thromboembolic events occurred despite standard doses thromboprophylaxis.^24^ Accordingly, high-dose LMWH treatment became standard in Swedish ICUs in April. We are still awaiting the results of ongoing RCT of LMWH at different doses (NCT04345848; NCT04344756)^25^, so we do not yet know the impact of high-dose LMWH on outcome. Corticosteroids became standard care for covid-19 patients requiring oxygen therapy after June 16, when the RECOVERY RCT showed survival benefits from corticosteroids.^26^ Prior to that, corticosteroids were predominantly used on the ICU, and their use increased over the study period (table 3). However, the infrequent use of corticosteroid treatment in March-May suggests that corticosteroids were unlikely to be a major cause for the decline in mortality among ICU patients.

Second, the high load of new admissions and of patients in hospital care for covid-19 in March and April (fig 2) may have contributed to the high initial mortality. This notion is supported by the result from healthcare region Stockholm-Gotland, which had the highest number of new covid-19 admissions/population in March-April (table E4) and also the clearest decline in mortality (fig E6).

Third, the decline in mortality may have been due to a change in the selection of covid-19 patients for hospital care. Among hospitalised covid-19 patients in the present study, the proportion admitted to ICU and receiving invasive mechanical ventilation decreased substantially over time (fig 6). In addition, the proportion of patients without comorbidity (CCI zero) increased significantly over time. These aspects indicate that the overall hospitalised covid-19 population was gradually less severely ill. This explanation is supported by an Italian study of covid-19 patients diagnosed in the emergency department between March and May, showing that the SARS-CoV-2 viral load in the upper respiratory tract gradually decreased and the proportion of patients requiring ICU care decreased over time.

^27^Fourth, the decline in mortality could perhaps be due to changes in virulence of the SARS-CoV-2 virus. Young et al.^28^ showed that a major deletion in the SARS-CoV-2 genome was associated with milder infection.^28^ Among 521 virus strains in Sweden with complete genome sequences, 19 different SARS-CoV-2 strain sequences were identified, 11 of which were identified in strains collected in March only.^29^ It will be important to investigate if the most prevalent SARS-CoV-2 strains in May and June had an inherent reduced virulence or had undergone genetic changes that reduced virulence.

The results of the present study, showing declining mortality over time in both non-ICU treated and ICU treated patients, combined with some previous studies showing declining mortality among ICU treated patients^8,12^, shed new light on covid-19 mortality and enables a more appropriate evaluation of the management of the pandemic. For instance, in studies using mortality as endpoint, the timing of inclusion may play a crucial role regarding outcome. The results of before-and-after studies on specific interventions should thus be interpreted with caution. The impact of an intervention during a high-mortality period does not necessarily apply to the same intervention during a period with significantly lower mortality. This is important when planning for allocation of healthcare resources to meet the next phases of the pandemic.

The present study has a number of strengths. First, it was a nationwide study including all hospitals in Sweden, minimising risk of selection bias. Second, it was based on national registers with standardised reporting on a national level, minimising bias of ascertainment. The unique national personal identification number of all Swedish residents enabled cross-linkage of registers at the individual level. Third, the inclusion of both non-ICU treated and ICU treated patients enabled analyses of the full nationwide cohort of hospitalised covid-19 patients. Fourth, the use of 60-day mortality provided a robust outcome measure with a follow-up long enough to enable hospital discharge of most patients.

The study also has limitations. First, clinical data regarding organ function were not available for non-ICU treated patients. Thus, it was not possible to determine degree of respiratory failure within the whole cohort. Second, data on do-not-resuscitate orders were not available and thus, it is not known if criteria for admission to ICU changed with time. However, SAPS3 scores on ICU admission were constant throughout the study period, indicating that the criteria for ICU admission had probably not changed considerably. Third, we did not have access to data on drug therapy on wards during hospital stay, and thus we could not appropriately assess the impact of different drug therapy for outcome. Fourth, the NPR lacks information from primary care, hence the CCI may be underestimated. However, replacing CCI with the individual comorbidities measured from both the NPR and the Prescribed Drug Register (thus catching primary care) in the models did not attenuate RR estimates further (not shown).

In conclusion, there was a distinct gradual decline in mortality for both non-ICU treated and ICU treated hospitalised covid-19 patients. Future studies are needed to address and explain this decline. The changing covid-19 mortality should be considered when the management and results of studies from the first pandemic wave are evaluated.

## Data Availability

The data underlying this article cannot be shared publicly due to regulations under the Swedish law. According to the Swedish Ethical Review Act, the General Data Protection Regulation, the Public Access to Information and Secrecy Act, data can only be made available, after legal review, for researchers who meet the criteria for access to this type of confidential data. Requests regarding the data may be made to the senior author.

https://www.socialstyrelsen.se/en/

## Contributors

All authors conceived and designed the study.

KS, EW, SW, ABB, MH, JH and HH acquired data.

EW and JH performed analyses and interpreted these together with KS, SW and HH.

EW and JH verified the underlying data in the article.

EW and JH drafted and finalised all tables and figures.

KS and EW contributed equally.

KS drafted the first version of the manuscript.

All authors had full access to data in the study and accept responsibility for submission for publication.

All authors revised the manuscript critically for important intellectual content and approved the final version for submission.

## Dissemination declaration

The results of this work will be disseminated to the public through press releases and media commentary. We are unable to directly provide the results of the research to study participants as this study analyzed deidentified data.

## Acknowledgments

This study was funded by Sweden’s National Board of Health and Welfare. The authors thank Anastasia Nyman for valuable discussion and support.

